# Prevalence and associated factors of advanced HIV disease among ART-naïve and ART-experienced people with HIV at AHF-supported sites in Zambia

**DOI:** 10.1101/2024.11.14.24317325

**Authors:** Webster Chewe, Chitalu Chanda, Benson M. Hamooya, Lukundo Siame, Ruth Lindizyani Mfune, Mazinga Kayembe, Eric Mpoyi, Nduduzo Dube

## Abstract

**Background:** Individuals with advanced HIV disease (AHD) have a heightened risk of mortality despite access to antiretroviral therapy (ART). Sub-Saharan Africa (SSA) has a disproportionate burden of AHD-associated deaths. However, there is limited country-specific data on AHD’s burden and associated risk factors to help design targeted interventions. Therefore, this study determined the prevalence and factors associated with AHD among ART-naïve and ART-experienced adults with HIV at five AIDS Healthcare Foundation (AHF)-supported facilities in Zambia.

**Methodology:** We conducted a cross-sectional study among 231 ART-naïve and ART-experienced people living with HIV (PLHIV) and collected demographic and clinical data using a structured questionnaire. The primary outcome was AHD, defined as a CD4 count less than 200 cells/µL using the VISITECT® CD4 point-of-care test. Multivariable logistic regression was used to assess factors associated with AHD.

**Results:** Among the study participants, 59.7% were female, and 54% were aged 19–35 years. Most participants were ART-naïve (79.2%), with 85.3% classified in WHO clinical stage 1. The prevalence of AHD was 47.6% (110/231), significantly higher among ART-naïve participants [51.9% (95/183)] compared to ART-experienced participants [31.2% (15/48), p=0.011]. ART-naïve participants aged 19–35 years exhibited a notably higher prevalence of AHD (51.6%) compared to those aged 13-18, 36-45, and >45 years (5.3%, 34.7%, and 8.4%, respectively) and had significantly higher baseline viral load values (25,000 copies/mL vs. 9,690 copies/mL, p=0.037). Additionally, among ART-naïve participants, a significantly higher proportion of individuals in WHO stages 2, 3, and 4 had AHD compared to those without AHD. Across all participants, the factors significantly associated with AHD included anemia (adjusted odds ratio [aOR] 3.03, 95% confidence interval [CI]: 1.15–7.97) and being from the New Masala Clinic health facility (aOR 3.79, 95% CI: 1.17–12.30)

**Conclusion:** Advanced HIV disease was prevalent, particularly among ART-naïve individuals, and it was significantly associated with anemia and health facility location. There is a need for formulation of interventions, especially among ART-naïve PLHIV who are anemic. Additionally, studies to integrate point-of-care diagnostics such as the VISITECT® CD4 test into routine HIV care be conducted in resource-limited settings are warranted for early detection of AHD.

## Introduction

Despite over three decades since the onset of the human immunodeficiency virus (HIV) pandemic, epidemic control remains elusive. Significant morbidity and mortality persist, particularly among individuals with advanced HIV disease (AHD) [1,2]. According to the World Health Organization (WHO), AHD is defined as CD4 count below 200 cells/µL or WHO clinical stage 3 or 4 [3,4]. The “test and treat” strategy has improved survival rates among people living with HIV (PLHIV), and the rollout of testing in most healthcare facilities has helped reach populations that were previously underserved [5,6]. As a result, treatment coverage has increased across many regions, with several countries reporting that over 95% of PLHIV are on antiretroviral therapy (ART) [7,8]. Furthermore, more than 70% of clients who initiate ART have CD4 counts above 200 cells/µL [9]. However, despite increased efforts in HIV screening and treatment, high mortality among PLHIV remains largely driven by and associated with AHD [10]. AHD is characterized by an increased vulnerability to malignancies and opportunistic infections such as bacterial infections, cryptococcal infections, and mycobacterial diseases like *Mycobacterium tuberculosis* and nontuberculous mycobacterium [11]. Additionally, AHD is often associated with anemia and weight loss [2]. Many PLHIV remain asymptomatic due to the natural progression of HIV infection, which often masks disease progression and leads to late presentation, especially in those with AHD [12,13].

Newly HIV-diagnosed individuals are among the most likely to present with AHD due to low rates of elite HIV control in low- and middle-income countries (LMICs) [14]. Without ART, most PLHIV present with unsuppressed HIV viral loads [15,16]. Furthermore, the risk of AHD is higher among those who are not on ART or have disengaged from care [17,18]. The prevalence of AHD is estimated to be 60% among those who have disengaged from care and 43.2% among ART-naïve PLHIV [19]. While the prevalence of AHD among ART-naïve PLHIV appears to be declining, there is limited trend analysis available for ART-exposed PLHIV who have access to care [20]. Approximately 60% of PLHIV on ART still have CD4 counts below 200 cells/µL, and this trend has remained static for years [21]. Disengagement from care continues to be a significant challenge in LMICs [1].

There is limited data on the prevalence and associated factors of AHD among ART-naïve PLHIV and those ART experienced in SSA information cardinal for policy formulation and in informing clinical practice. Therefore, this study aimed to estimate the prevalence and factors associated with AHD among PLHIV who are ART-naïve and ART-experienced in five AIDS Healthcare Foundation (AHF)-supported health facilities in Zambia.

## Methods

### Study site and study design

A multicenter, cross-sectional study was conducted among individuals with HIV in five health facilities across Zambia from January 8 to May 31, 2024. The participating sites included AHF stand-alone ART clinics at Chifundo and Lusungu in Lusaka District, Kapiri Mposhi Urban Clinic (Kapiri Mposhi District), New Masala Clinic (Ndola District), and Shampande Clinic (Choma District). These facilities were purposively selected to represent diverse regions across four provinces of Zambia.

### Sample size and sampling technique

The sample size of 231 participants was determined based on the available testing resources at the selected five facilities during the study period. Purposive sampling was employed to include all patients who met the study’s inclusion criteria and were present at the facilities at the time of data collection.

### Study Definitions

Advanced HIV disease (AHD) was defined as any person living with HIV who had a CD4 count of <200 cells/µL or was categorized under WHO clinical stage 3 or 4. HIV status was determined according to Zambia’s national standards [4,22].

Antiretroviral therapy (ART) status was classified as either ART-naïve or ART-experienced. ART-naïve referred to any person living with HIV who had never received ART, while ART-experienced referred to those who had previously received or were currently receiving ART. Anaemia was defined as a hemoglobin concentration of less than 13.0 g/dL in males and less than 12.0 g/dL in females [23].

HIV Viral load was defined as a measure of the amount of HIV RNA (viral particles) in a participant’s blood expressed in copies of the virus per milliliter of blood (copies/mL). Suppressed HIV viral load was defined as a viral load of fewer than 200 copies/mL. Low-level viremia (LLV) was defined as a viral load between 200 – 999 copies/mL. Unsuppressed HIV viral load was defined as a viral load of greater or equal to 1,000 copies/mL [24].

Silent transfer was defined as the situation where a person living with HIV presents to a new healthcare facility with a viral load ≤1000 RNA copies/mL, without prior formal documentation or referral from their previous facility [25].

### Data and sample collection

Baseline data were collected using a structured questionnaire developed and administered by trained research assistants. The data included demographic (age and sex) and clinical (ART status, ART regimen, WHO clinical staging, hemoglobin levels, VISITECT® CD4 test, viral load, and urine LAM test) characteristics. Approximately 8 mL of venous blood was collected from each consenting participant into an EDTA tube and stored at room temperature until transported to the designated site laboratory for analysis of hemoglobin levels, CD4 count, and viral load determination. Blood samples were analyzed within 24 hours, and CD4 cell counts were measured using the VISITECT® CD4 test according to the manufacturer’s specifications.

### Data analysis plan

Data were cleaned in Microsoft Excel and analyzed using STATA 15. Descriptive statistics were used to understand the distribution of variables among the study participants. The Shapiro Wilk test was used to determine the normality of a continuous variable, and since there was no normality, we used the Wilcoxon rank sum test to ascertain a statistical difference between the two medians. A relationship between two categorical variables was determined using the Pearson chi-square test or Fisher’s exact test, depending on applicability. Multivariable logistic regression was used to ascertain factors associated with AHD. Factors in the final model were selected based on the previous literature. A p-value < 5% was considered statistically significant.

### Ethics

The study protocol was reviewed and approved by the Tropical Diseases Research Centre (TDRC) ethics review committee (TRC/C4/12/2023) and The National Health Research Authority (NHREB006/29/12/2023). Permission was obtained from the provincial, district, and facility in-charges where the study took place. Written informed consent was obtained from study participants. Data were collected without personal identifiers, kept secure and confidential and were applicable, only available to the principal and co-principal investigators.

## Results

### Basic characteristics of the study participants

This study included 231 participants, of whom 59.7% (138/231) were female, and 54% (125/231) were in the age group of 19-35 years. Most patients were ART-naïve (79.2%, 183/231), while 20.8% (48/231) had prior ART experience. The majority of participants (85.3%, 197/231) were classified as being in WHO clinical stage 1, and 5.9% (3/51) had positive urine LAM tests. Additionally, about one-third of the participants were anemic (33.5%, 77/231).

### The burden of advanced HIV disease in ART-naïve and ART-experienced Participants

The overall prevalence of AHD was 47.6% (110/231), higher among ART-naïve participants (51.9%, 95/183) than ART-experienced participants (31.2%, 15/48), p=0.011, as shown in Fig 1.

**Figure 1.**
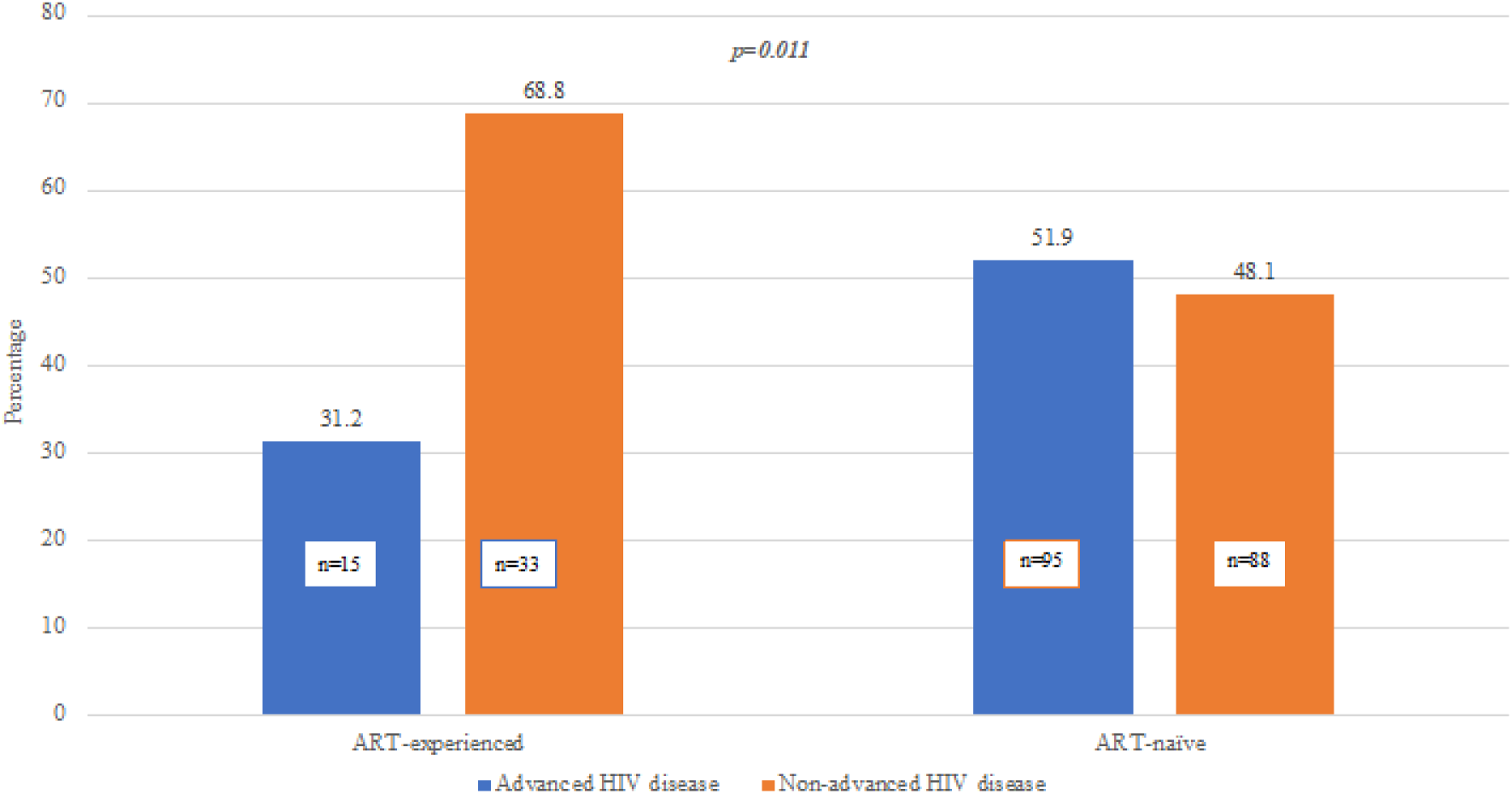
Prevalence of Advanced HIV disease and Non-Advanced HIV disease among ART-naïve and ART-experienced Participants

### Relationship between AHD and other study variables

Among ART-naïve participants with AHD, those in the 19–35 age group had a higher prevalence of AHD (51.6% vs. 5.3%, 34.7%, and 8.4%) and were more likely to be in WHO HIV Clinical Stage 1 (80% vs. 13.7% and 3.2%). ART-naïve individuals with AHD also had significantly higher baseline viral load values, 25,000 vs. 9,690, p = 0.037. In the overall analysis (combining ART-experienced and ART-naïve participants), those with AHD had higher viral load values at enrolment compared to those without AHD, 26,950 vs. 5,090, p = 0.012. Additionally, anemia was more prevalent in participants with AHD than in those without (41.8% vs. 25.8%, p = 0.010), see Table 1

**Table 1:**
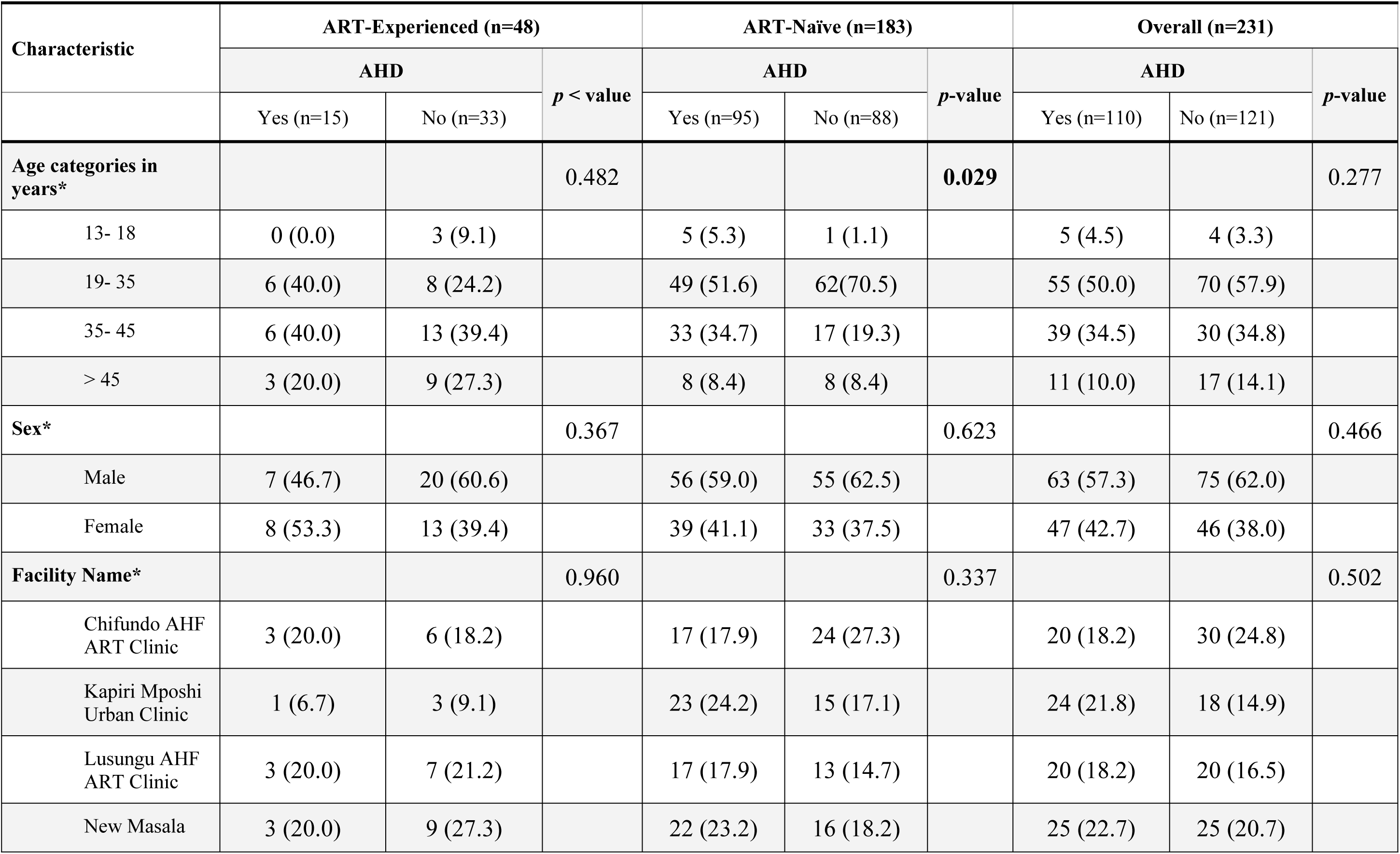
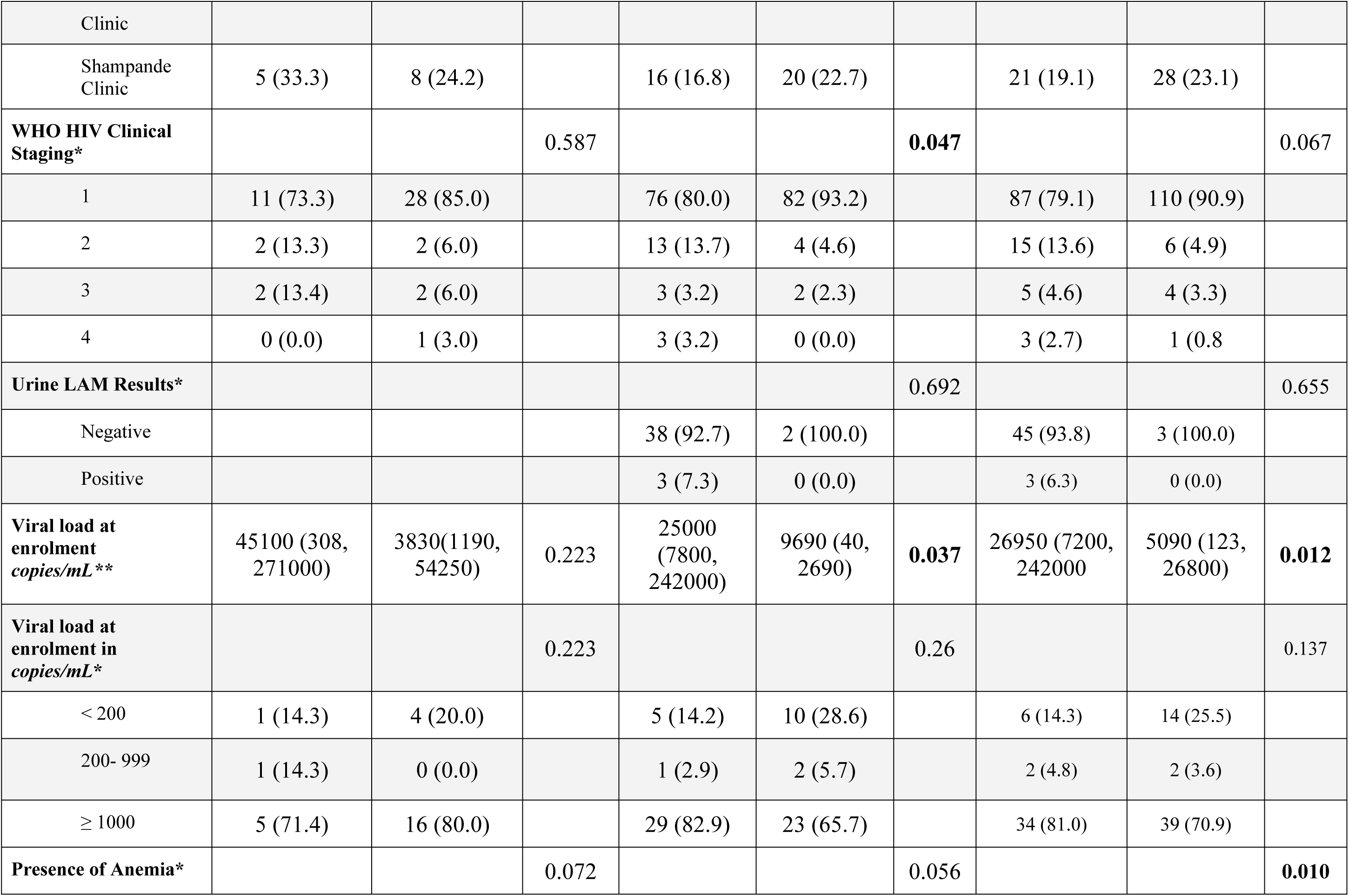
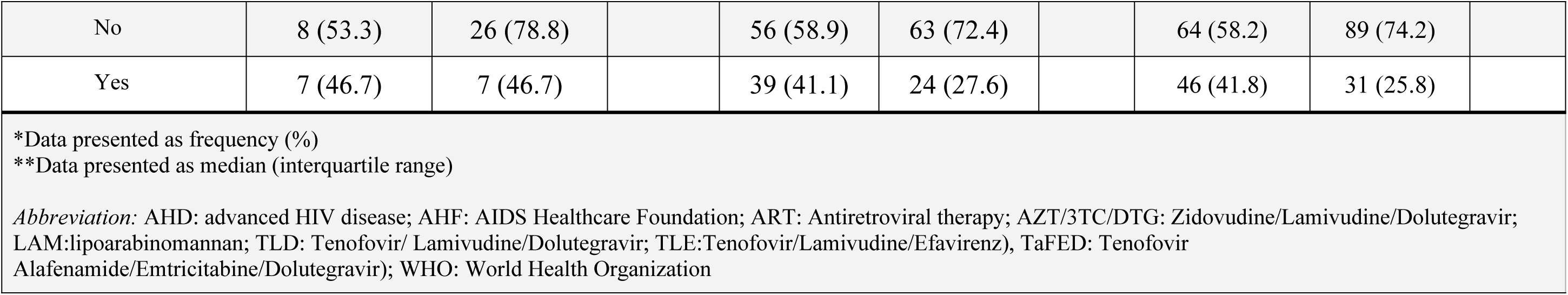
Demographic and Clinical Factors sorted according to advanced HIV disease among ART Experienced and Naïve Participants.

#### Regression Analysis of Factors Associated with AHD among ART-Naïve, ART-Experienced, and Combined Group Participants

Table 2 presents the results of bivariable and multivariable analyses assessing factors associated with AHD. In the overall crude analysis (combining ART-experienced and ART-naïve participants), individuals with anemia had 2.06 times higher odds of being diagnosed with AHD compared to non-anemic individuals (crude odds ratio [cOR] = 2.06; 95% CI: 1.18–3.60; p = 0.011). Participants with WHO clinical stage 2 had 3.16 times higher odds of AHD diagnosis compared to those not in Stage 2 (cOR = 3.16; 95% CI: 1.17–8.49; p = 0.022). In the adjusted analysis, participants receiving care at New Masala Clinic had significantly higher odds of being diagnosed with AHD compared to those from Lusungu clinic (adjusted odds ratio [aOR] = 3.79; 95% CI: 1.17–12.30; p = 0.027). Additionally, participants with anemia had 3.03 times higher odds of receiving an AHD diagnosis compared to non-anemic participants (aOR = 3.03; 95% CI: 1.15–7.97; p = 0.024).

**Table 2.** Logistic Regression Analysis of Factors Associated with Advanced HIV Disease among ART Experienced and Naïve Participants.

| Variable | ART-Experienced (n=48) |  |  |  | ART-Naïve (n=183) |  |  |  | Overall (n=231) |  |  |  |
| --- | --- | --- | --- | --- | --- | --- | --- | --- | --- | --- | --- | --- |
|  | Crude analysis |  | Adjusted analysis |  | Crude analysis |  | Adjusted analysis |  | Crude analysis |  | Adjusted analysis |  |
|  | cOR (95% CI) | p-value | aOR (95% CI) | p-value | cOR (95% CI) | p-value | aOR (95% CI) | p-value | cOR (95% CI) | p-value | aOR (95% CI) | p-value |
| **Age categories in years** |  |  |  |  |  |  |  |  |  |  |  |  |
| 13 - 18 |  |  |  |  | 5.0 (0.47, 52.96) | 0.181 | - | - | 1.93 (0.42, 8.81) | 0.395 | 9.35 (0.77, 112.6) | 0.078 |
| 19 - 35 | 2.25 (0.41, 12.09) | 0.345 |  |  | 0.77 (0.27, 2.21) | 0.663 | 0.71(0.08, 5.83) | 0.755 | 1.21 (0.52, 2.80) | 0.649 | 1.75 (0.42, 7.21) | 0.441 |
| 36 - 45 | 1.38 (0.27,7.04 ) | 0.695 |  |  | 1.94 (0.62, 6.08) | 0.255 | 0.87(0.08, 5.83) | 0.908 | 2.00 (0.82, 4.91) | 0.127 | 1.25 (0.28, 5.61) | 0.770 |
| > 45 | ref |  |  |  | Ref |  | Ref |  | Ref |  | Ref |  |
| **Sex** |  |  |  |  |  |  |  |  |  |  |  |  |
| Male | Ref |  |  |  | Ref |  | Ref |  | Ref |  | Ref |  |
| Female | 0.57 (0.17, 1.94) | 0.36<br>9 |  |  | 0.84 (0.46, 1.53) | 0.582 | 0.49 (0.12, 1.95) | 0.319 | 0.82 (0.48, 1.39) | 0.466 | 0.68 (0.23, 2.00) | 0.487 |
| **Facility Name** |  |  |  |  |  |  |  |  |  |  |  |  |
| Lusungu AHF ART Clinic | Ref |  |  |  | Ref |  | Ref |  | Ref |  | Ref |  |
| Chifundo AHF ART Clinic | 1.17 (0.16, 8.09) | 0.87<br>6 |  |  | 0.54 (0.20, 1.40) | 0.207 | - | - | 0.67 (0.28, 1.54) | 0.344 | - | - |
| Shampande Clinic | 1.45 (0.25, 8.42) | 0.67<br>3 |  |  | 0.62 (0.23, 1.62) | 0.324 | - | - | 0.75 (0.32, 1.73) | 0.502 | - | - |
| Kapiri Mposhi Urban Clinic | 0.77 (0.06, 10.86) | 0.85<br>2 |  |  | 1.12 (0.42, 2.98) | 0.818 | 8.82 (0.88, 88.23) | 0.064 | 1.33 (0.56, 3.18) | 0.517 | 3.50 (0.74, 16.67) | 0.115 |
| New Masala Clinic | 0.78 (0.11, 5.10) | 0.79<br>3 |  |  | 1.04 (0.39, 2.76) | 0.919 | 4.67(1.05, 20.74) | **0.043** | 1.00(0.43, 2.29) | 1.00 | 3.79 (1.17, 12.30) | **0.027** |
| **Viral load at enrolment in copies/mL** |  |  |  |  |  |  |  |  |  |  |  |  |
| < 200 |  |  |  |  | Ref |  | Ref |  | Ref |  | Ref |  |
| 200 - 999 |  |  |  |  | 1.00 (0.07, 13.86) | 1.000 | 0.63 (0.03, 10.36) | 0.749 | 2.33 (0.26, 20.65) | 0.446 | 2.48 (0.21, 28.71) | 0.466 |
| ≥ 1000 |  |  |  |  | 2.43 (0.72, 8.13) | 0.148 | 2.12 (0.39, 11.46) | 0.382 | 2.03 (0.70, 5.88) | 0.190 | 1.91 (0.53, 6.85) | 0.317 |
| **WHO HIV Clinical Staging** |  |  |  |  |  |  |  |  |  |  |  |  |
| 1 | Ref |  |  |  | Ref |  | Ref |  | Ref |  | Ref |  |
| 2 | 2.55(0.32, 20.38) | 0.379 |  |  | 3.55 (1.10, 11.38) | **0.033** | 2.67 (0.30, 23.76) | 0.379 | 3.16 (1.17, 8.49) | **0.022** | 2.97 (0.58, 15.15) | 0.190 |
| 3 |  |  |  |  | 1.64 (0.27, 10.08) | 0.593 | - | - | 1.58 (0.41, 6.06) | 0.505 | 0.07 (0.003, 1.32) | 0.076 |
| 4 |  |  |  |  | - | - | - | - | 3.79 (0.38, 37.10) | 0.252 | - | - |
| **Presence of Anemia** |  |  |  |  |  |  |  |  |  |  |  |  |
| No | Ref |  |  |  | Ref |  | Ref |  | Ref |  | Ref |  |
| Yes | 3.25 (0.87, 12.08) | 0.079 |  |  | 1.86 (0.99, 3.47) | 0.051 | 2.92 (0.90, 9.46) | 0.073 | 2.06 (1.18, 3.60) | **0.011** | 3.03 (1.15, 7.97) | **0.024** |
| *Abbreviations:* AHD: advanced HIV disease; AHF: AIDS Healthcare Foundation; aOR: adjusted odds ratio; ART: antiretroviral therapy; cOR: crude odds ratio; Ref: reference group; WHO: World Health Organization |  |  |  |  |  |  |  |  |  |  |  |  |

In the crude analysis among ART-naïve participants, those from Masala clinic were 4.67 times more likely to be diagnosed with AHD compared to those from Lusungu clinic (cOR = 4.67; 95% CI: 1.05–20.74; p = 0.043). Participants with WHO clinical stage 2 had 3.55 times higher odds of AHD diagnosis compared to those in WHO stage 1 (cOR = 3.55; 95% CI: 1.10–11.38; p = 0.033). In the adjusted analysis, participants from New Masala Clinic had significantly higher odds of being diagnosed with AHD compared to those from Lusungu clinic (aOR = 4.67; 95% CI: 1.05–20.73; p = 0.043).

## Discussion

The overall prevalence of AHD in our study was 47.6%, with a notably higher prevalence among ART-naïve individuals (51.9%) compared to ART-experienced individuals (31.2%). This finding is consistent with studies from other regions, such as South Africa and Sierra Leone, which also highlight the persistent challenge of AHD despite the availability of ART [11,19]. For instance, a study in South Africa reported a higher prevalence of AHD among ART-experienced individuals (58.6%) compared to ART-naïve individuals (43.45%) [19]. Similarly, in Sierra Leone, AHD prevalence was found to be 58.6% among ART-experienced individuals, higher than the 43.5% prevalence among ART-naïve youth aged 15-24 years [11]. These findings from South Africa and Sierra Leone underscore the complexity of AHD management, with ART-experienced individuals facing significant challenges despite prolonged ART use [9,11,12,19,26,27].

This high prevalence, particularly among ART-naïve patients, underscores the need for targeted community innovations to ensure that The Joint United Nations Programme on HIV and AIDS (UNAIDS) first 95 target is globally met and that case finding efforts for HIV are comprehensive [28,29]. The significant burden of AHD among both ART-naïve and ART-experienced patients could be due to the lack of targeted HIV case finding, particularly in certain age groups, despite achieving 95% of patients knowing their HIV status and 95% being on ART [30].

A notable finding of this study was the significant association between anemia and AHD, as observed in both ART-naïve and ART-experienced participants. This association is consistent with existing literature, where anemia is a common comorbidity among PLHIV, particularly those with advanced immunosuppression [31]. The presence of anemia in this population is multifactorial, resulting from chronic inflammation, opportunistic infections, and ART-related toxicities [32,33]. The study further found that most participants classified in WHO Stage 1 did not have AHD, which aligns with expected clinical staging outcomes for AHD. In contrast, a higher prevalence of AHD was observed among participants in WHO stages 2, 3, and 4, indicating that advanced clinical staging correlates with the CD4-defined criterion for AHD. This alignment supports the validity of both WHO staging and CD4 counts in identifying individuals with advanced immunosuppression [34–36]. Additionally, the study identified positive results for tuberculosis screening even among participants in WHO Stage 1, suggesting that point-of-care diagnostics like the VISITECT® CD4 test may offer greater sensitivity in detecting AHD. This finding underscores the potential benefit of integrating rapid diagnostic tools into routine clinical practice to improve early identification of patients at risk for poor outcomes, thereby supporting more timely interventions.

The 19–35-year age group had a higher proportion of AHD among ART-naïve participants. This finding aligns with other studies that have reported a higher prevalence of AHD among youths [11]. The recent Zambia population-based HIV impact assessment (ZAMPHIA) report also highlights the higher incidence of HIV in this age group, coupled with challenges in accessing HIV preventive strategies [30]. These findings may reflect differences in health-seeking behaviors, adherence patterns, or immunological responses among PLWH [37,38]. Therefore, there is a need for tailored interventions targeting specific demographic groups that could help address these disparities.

The study found no significant association between sex and AHD, indicating that both male and female patients are equally at risk of presenting with advanced disease, which could be due to the smaller sample size of our study. This result contrasts with findings from other studies that suggest female sex may be protective [11,26].

Another noteworthy finding was that 14.3% of both ART-naïve and ART-experienced individuals with a viral load of less than 200 copies/mL presented with AHD. Concurrent AHD and viral load suppression were observed in 14.3% of ART-experienced and 14.1% of ART-naïve patients. This phenomenon may be attributed to “HIV elite control” or “silent transfer,” where patients achieve viral suppression despite presenting with AHD [39]. These results were higher than those observed in the Zimbabwe Population-Based HIV Impact Assessment (ZIMPHIA) study, which reported 13% of patients with concurrent viral load suppression and AHD [40]. Additionally, low-level viremia, defined as viral loads between 200 and 1000 copies/mL, was associated with AHD, although this association did not reach statistical significance. While the current study did not find a significant association between viral load and AHD, further investigation is warranted into the potential effects of low-level viremia on AHD outcomes. Previous studies have linked low-level viremia to adverse health outcomes, although the association was not statistically significant in the present analysis, possibly due to the small sample size [41].

The significant burden of AHD and the high prevalence of anemia observed in this study underscore the need for comprehensive care strategies that extend beyond ART provision [42]. Enhancing AHD screening through point-of-care diagnostics, along with further research on the impact of managing comorbidities like anemia, is essential to improve AHD outcomes.

### Study limitations and strengths

The study has several limitations. First, the cross-sectional design limits the ability to establish causality between observed factors and AHD. Second, the study relied on a small sample size which may not be representative of the broader population of PLHIV across the country. Third, the use of novel diagnostic methods for AHD, such as the VISITECT® CD4 Advanced Disease Test could lead to variability in prevalence estimates. Despite the limitations, the study was able to quantify the burden and factors associated with AHD information valuable for informing policy and clinical practice.

## Data Availability

No restriction

## Conclusion

This study underscores the significant burden of AHD among both ART-naïve and ART-experienced participants at selected AHF-supported facilities in Zambia. The high prevalence of AHD, particularly among those newly diagnosed, highlights the ongoing challenges in early HIV diagnosis and the need for targeted intervention, especially considering that the first 95% has been achieved by Zambia. Anemia and health facility location were identified as factors significantly associated with AHD. These findings emphasize the urgent need for comprehensive care strategies that integrate routine anemia screening alongside HIV treatment. Additionally, the use of point-of-care diagnostics, such as the VISITECT® CD4 test, proved valuable for detecting AHD, particularly in resource-limited settings. The observed prevalence of AHD in patients with suppressed HIV viral loads warrants further investigation, as it may suggest underlying factors such as HIV elite control or “silent transfer”. Further exploration into the immunology of elite HIV control and potential vaccine implications is needed.

## Authors Contributions

Conceptualization: W.C., and C.C. Writing (original draft): W.C., C.C., and B.M.H. Writing (review and editing): W.C., C.C., B.M.H., L.S., R.L.M., M.K., E.M., and N.D. Visualization: W.C. Data collection: W.C., C.C., M.K., and E.M. Data analysis: W.C., C.C., B.M.H, and L.S. Supervision: C.C., and E.M. All authors have read and agreed to the published version of the manuscript.

## Acknowledgment

The authors take the opportunity to thank the AIDS Healthcare Foundation Zambia, Zambia Ministry of Health and Management of individual health facilities that enabled us to collect the data for this study.

